# Simple quantitative assessment of the outdoor versus indoor airborne transmission of viruses and covid-19

**DOI:** 10.1101/2020.12.30.20249058

**Authors:** B.R. Rowe, A. Canosa, J.M. Drouffe, J.B.A. Mitchell

**Affiliations:** Rowe Consulting, 22 chemin des moines, 22750 Saint Jacut de la Mer (France); CNRS, IPR (Institut de Physique de Rennes)-UMR 6251, Université de Rennes, 35000 Rennes, (France); 31B Chemin du Couvent, 91190 Gif-sur-Yvette (France); MERL-Consulting SAS, 21 Rue Sergent Guihard, 35000 Rennes (France)

**Keywords:** Covid-19, virus airborne transmission, outdoor versus indoor transmission, quantitative risk assessment, meteorological and geographical influence

## Abstract

In this paper we develop a simple model of the inhaled flow rate of aerosol particles of respiratory origin *i.e*. that have been exhaled by other humans. A connection is made between the exposure dose and the probability of developing an airborne disease. This allows a simple assessment of the outdoor versus indoor risk of contamination to be made in a variety of meteorological situations. It is shown quantitatively that for most cases, the outdoor risk is orders of magnitude less than the indoor risk and that it can become comparable only for extremely specific meteorological and geographical situations. It sheds a light on various observations of Covid-19 spreading in mountain valleys with temperature inversions while at the same time other areas are much less impacted.

**Highlights:**

- Risk of covid-19 airborne transmission.
- Quantitative assessment of outdoor versus indoor airborne risk of transmission.
- Meteorological and geographical influence on covid-19 airborne transmission.

## 1 Introduction

The purpose of the present paper is to develop a simple quantitative assessment of the relative risk between indoor and outdoor environments for the so-called “aerosol” or “airborne” transmission of viruses and for different outdoor situations. The goal is to assist in public health policy and recommendations.

Respiratory diseases represent a serious burden for global public health. It should be remembered that in western countries, prior to the advent of antibiotic drugs they were the primary cause of death. Antibiotics, however, are essentially inactive for virus borne illnesses, except in their ability to prevent secondary infection. In the case of mutating viruses, vaccines have to evolve constantly as in the case of influenza viruses. Therefore, it is essential to understand the problem of virus transmission in order to provide effective guidance for the mitigation of epidemics.

Amongst respiratory diseases and according to the World Health Organization, influenza which is caused by viruses of various kinds, each year leads to the premature death of between 290,000 and 650,000 people (WHO, 2017) and measles more than 140 000 (WHO, 2019b). At the beginning of the 21^st^ century, new respiratory viruses have appeared such as the SARS-COV-1 in 2003, a coronavirus that emerged first in China in 2002 and caused severe respiratory disease leading often to pneumonia with a rather high (around 10%) mortality. Fortunately, the spread of that epidemic was limited mainly to Asia and came to an end before the end of 2003 (CDC, 2017). Therefore, despite its seriousness, the total mortality of SARS-COV-1 remained low. More recently in 2009, the H1N1 pdm2009 flu virus emerged, and it has been estimated by the CDC (US Center of Disease Control) that during its first year of circulation it killed 0.001 to 0.007 % of the world population (CDC, 2020). It has been circulating since, causing significant health problems in various countries.

For comparison previous epidemics in the last century included the 1968 H3N2 flu which killed around 0.03% of the world population (CDC, 2019b), and the 1918 H1N1 pandemic (so called Spanish flu) had a much more terrible impact ranging from 1 to 3% (CDC, 2019a).

The actual pandemic linked to the new SARS-COV2 coronavirus which emerged in China at the end of 2019, has already resulted in mortality close to 0.02% of the world population (JHU, 2020). The illness caused by this virus has been named COVID-19 for COronavirus Virus Disease of 2019 and has led many governments to take stiff measures such as lockdowns with severe damage inflicted on the economy and secondary effects on health. Therefore, and as stated above, a good knowledge of the actual transmission routes is essential in order to take rational and scientifically based decisions to mitigate virus spread without destroying social life and the economy.

It is commonly admitted that respiratory viruses are transmitted in three ways. The first is via “direct contact”: it means that an infected person can transmit a given amount of virus to a person in close contact, either by sneezing or coughing and even talking and breathing, thus emitting a variety of micro-droplets that can be projected directly onto the mucosa (lips, nose and eyes) of this person, or onto the skin and clothes and subsequently transmitted by the hands to the mucosa. The second way is linked to objects that have been contaminated in the same way and referred to in medical science as “fomites”: it is then expected that, even without direct contact with the infected person, touching the contaminated object with bare hands can lead to contamination. The third route involves a persistent aerosol formed by the smallest particles emitted by an infected person that can subsequently be breathed in. This is known as “airborne transmission” or “aerosol transmission”.

At the beginning of the COVID-19 pandemic, airborne transmission was minimized, if not outright denied, by health authorities either by the WHO or by governmental agencies in a variety of countries, such as the CDC (Center of Disease Control) in the US or the HAS in France (Haute Autorité de Santé). Therefore, the recommendations for mitigation of the epidemic were mainly based on the first two methods of transmission: social distancing which means not coming into close contact with someone else (with a recommended distance between one and two meters as defined in different countries), together with frequent washing and disinfecting hands and surfaces. Finally, in addition to these preventative measures for individuals, a mitigation strategy of testing-tracing-isolating that may harm privacy and promote digital surveillance, based on new digital tools and tests was often adopted (Rowe *et al*., 2020).

However, several reasons, based as much on scientific work as on observation, cast great doubt on the fact that aerosol transmission could be negligible. In fact, it appears now that it could be a major way of transmission in addition to close contact. The key role for fomites itself is now contested (Goldman, 2020).

Microdroplets that move in the air experience a drag force F_D_ that results in a terminal velocity. For small particles, the drag force follows the well-known Stokes law (Stokes, 1851) and is proportional to the radius of the particle:

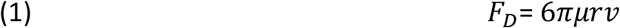

where *μ* is the air viscosity, with *r* and *v* respectively being the radius and velocity of the particle.

The force of gravity experienced by the particle is proportional to its mass *i.e*. to the third power of the radius, hence it is understandable that, below a given size, particles can remain a very long time in air (as fog) and even nearly indefinitely due to the natural and relentless movement of atmosphere, whether indoors or outdoors.

For a relative humidity lower than 100% the microdroplet can evaporate in a very short time, with a reduction in volume of an order of magnitude or more (Morawska, 2006; Nicas *et al*., 2005; Vejerano and Marr, 2018) due to water loss. This can lead to the formation of dry nuclei with a high biological load including viruses. Due to their size reduction these particles will remain airborne and represent a serious infectious hazard.

These phenomena were recognized and developed in a visionary paper by Wells as soon as 1934 which concluded that they result in contaminated air which can lead to the contagion of persons just by respiration (Wells, 1934). This third route of contamination is well recognized in a variety of disease such as measles (WHO, 2019a).

Due to the importance of respiratory diseases, researchers have not awaited COVID-19 to study and characterize aerosols emitted by human (Morawska *et al*., 2009) either by simple respiration or coughing, sneezing, talking, and singing. The behavior of the emitted aerosols (Bourouiba *et al*., 2014; Bourouiba, 2020) and the problem of their infectiousness (Buonanno *et al*., 2020) has also been studied. In fact, the scientific community has been the first to rise a cry of alarm regarding the most probable role of aerosol transmission (Borsellino *et al*., 2020; Morawska and Cao, 2020; Morawska and Milton, 2020).

However, it is mainly observations that lead to the conclusion that airborne transmission is a key route of contamination of COVID-19. Several situations have been reported involving high contamination rates at a given location, known as “super-spreader” or “cluster” events. An exceptionally large majority of these are indoor events which, as we shall see, is consistent with airborne transmission. These include contamination on cruise ships (Azimi *et al*., 2020), public transportation (Yang *et al*., 2020), restaurants (Lu and Yang, 2020), religious ceremonies (James *et al*., 2020) amongst others. The key role of aerosol transmission in these events has been discussed in a recent paper (Shen *et al*., 2020).

The seasonality of influenza is well known (Lofgren *et al*., 2007; Tamerius *et al*., 2013) and it is now largely admitted in the case of COVID-19 (Mattiuzzi *et al*., 2020). The importance of atmospheric parameters such as temperature and humidity are also well recognized in both cases (Marr *et al*., 2019; Pica and Bouvier, 2012). The strong correlation with weather and climate could be partly due to physiological reasons (Eccles, 2002; Rahman and Williams, 2021) but is a strong argument for the aerosol route.

Other arguments could come from other observations: the occurrence of straightforward pneumonia in some patients without any symptoms in the upper respiratory tract is completely consistent with the inhalation of infective microdroplets directly into the lung (Karimzadeh *et al*., 2020; Yezli and Otter, 2011). A difference has also been observed between the occurrence of severe forms of the disease between males and females (Peckham *et al*., 2020), with fatal outcomes being more probable for men. Apart from other physiological reasons, which are beyond the scope of this paper, we suggest that the difference between male and female respiration, males having a much deeper inspiration (LoMauro and Aliverti, 2018) together with aerosol transmission, could at least partly explain this difference.

In the present paper simple calculations and arguments lead to a remarkably simple formula for quantifying the relative level of exposure to the disease (which will be defined in section 3) and the relative probability of developing a disease between indoor and outdoor situations. Of course outdoor meteorological parameters are essential for quantitative assessments, a problem closely related to the science of air pollution.

Outdoors, only special situations of temperature inversion could lead to situations with an outdoor risk comparable to that found indoors. The discussion sheds light on the climatic and weather correlations that have been observed with the spread of the disease.

The present paper is organized as follow: in section 2 a short review is done of what is known about human exhaled droplets and aerosols. Then in section 3 modeling indoor situation is discussed and a simple calculation of the level of exposure (together with its definition) is presented. Note that the deep current knowledge of human aerosol reviewed very briefly in section 2 is not needed nor used for the sake of simplicity in section 3. In section 4 an airshed calculation of the outdoor level of exposure is described. Here the main unknown is the airshed “height” which is discussed at length in the supplementary materials (hereafter SM). The respective relative level of exposure and of disease probability between outdoor and indoor situations are discussed in section 5 with numerical applications and a general discussion. Section 6 deals of atmospheric markers of risk outdoor and indoor.

The main result of the present work, highlighted in section 5 and in the conclusion (section 7) is that the outdoor risk, except in special meteorological situations with very stable atmosphere and exceptionally low wind, is generally far lower than indoor risk (often by orders of magnitudes) and that indoors, **the fresh air ventilation rate is a key factor to mitigate the risk**. This last point has been pointed out by several researchers (Gao *et al*., 2016; Morawska *et al*., 2020) and is now recognized by the authorities.

## 2 Human emissions

### 2.1 Human respiratory characteristics

The first feature of human respiration is that we inhale fresh air mainly composed of nitrogen (78%) and oxygen (21%) and of various minor species including carbon dioxide (0.04%), natural aerosols and possible pollutants. In the exhaled air the concentration of carbon dioxide is enhanced to a much larger value, typically about 4-5% (Neronov *et al*., 2017). It also contains a variety of microdroplets of various sizes that come from the respiratory tract and are mainly composed of water (98.2%) (Chen and Zhao, 2010). As discussed in the introduction and in the SM the largest microdroplets fall to the ground over a relatively short distance. Hence the recommendation of social distancing of one to two meters. However, the smallest particles are able to stay in suspension in the air leading to the creation of a “human” aerosol. It has been commonly admitted that the border between the two cases is defined for a radius of 5 micrometers (Gralton *et al*., 2011) although such a simple discrimination has been largely disputed and that a variety of “borders” can be found in the literature (Bourouiba *et al*., 2014; Bourouiba, 2020; Morawska, 2006). The mean value at rest of the exhaled (inhaled) volume for adult is 500 ml (Tortora and Derrickson, 2016) with a normal frequency of 9-12 cycles per minute (Barrett *et al*., 2012) which leads to a mean air flow rate of around 5-6 l/mn that is used in the present work. Note that with higher physical activity the frequency hardly changes, and the higher flow rate comes from a higher inhaled/exhaled volume.

These water-based microdroplets contain mucus and possible viruses and bacteria, hence their potential role in contamination. This fact has led various researchers to study the physical and biological characteristics of emitted microdroplets. Clearly a study of their size distribution is fundamental to know if their aerosolization is possible, leading then to this route of contamination by an infected people.

As far back as 1945, Duguid conducted experiments using impact of exhaled microdroplets on celluloid slide followed by micrometry. His results were published in a seminal paper (Duguid, 1945), however in those days, most of the very powerful modern in-situ particle size analyzing methods did not exist. Since then, numerous studies have been performed (see for example (Alsved *et al*., 2020; Yang *et al*., 2007). Amongst them, the most powerful facility dedicated to this problem has been built at the Queensland University of Technology, at Brisbane, in Australia. A special wind tunnel (Morawska *et al*., 2009) allows a human emitter to be isolated in completely clean air (*i.e*., aerosol particles in the air are removed prior to experiment) and a variety of particle size analyzers are used to derive the complete particle size distribution. The research team led by L. Morawska has published several papers (Johnson and Morawska, 2009; Johnson *et al*., 2011; Morawska *et al*., 2009) amongst others which provide an overview of size distribution exhaled for a variety of human activities from breathing to coughing. Results clearly show that humans emit many particles that are aerosolized in different size modes associated with distinct processes: one occurring deeper or less in the respiratory tract.

Another physical parameter of exhaled air is its temperature which is lower than the human body temperature but can still be much higher than the ambient temperature, especially in wintertime at mid latitudes. Values of around 32-34°C have been widely reported (Carpagnano *et al*., 2017). Therefore, emitted puffs of air can rise by buoyancy in colder air.

In addition to these physical aspects, it is also important to characterize the emitted aerosol from a biological point of view. The coronavirus content in the fluid of respiratory tracts of infected people has been studied (To *et al*., 2020; Zou *et al*., 2020). However, such studies do not allow a quantitative estimation of aerosol infectivity to be deduced. One of the most used models in aerosol contamination is that of Wells-Riley which uses the concept of a quantum of infection and is described in the next section. The quantum of infection rate of production per infected people is subsequently determined by epidemiological observations. A more recent concept is the Minimum Infective Dose (hereafter MID) which can be defined as the minimum dose of viruses that can initiate infection in a given proportion of receptors. The factors influencing this dose are important for the development of any risk assessment. MID estimates are often determined by infecting young, healthy volunteers, which is of course restricted to non-dangerous viruses like those responsible of common cold (Yezli and Otter, 2011). The value of the MID is influenced by a variety of factor such as the route of inoculation, vulnerability of volunteers etc. Therefore, the links between the MID and the quantum of infection is not straightforward (Jones and Su, 2015; Sze To and Chao, 2010; Yezli and Otter, 2011). Although details of these biological and medical characteristics of disease transmission are beyond the field of competences of the present authors, what can be retained is that it is widely recognized that the virus dose received by a receptor is the main parameter of disease transmission. This justifies the quantification of relative risk using concepts based on level of exposure which are developed in section 3, as long as comparable situations are considered.

### 2.2 Behavior of the emitted aerosol

Many of the recommendations of the WHO and government agencies have been based on an analysis of the dynamical behavior of a single particle in still air. However, an aerosol is in fact a two-phase medium (gas plus particles) with a much more complicated behavior as is well known in the physics of atmospheric pollution. Several researchers have therefore been interested in the description of the air flow emitted by a person, either as gas (Gupta *et al*., 2010) or as aerosols (Bourouiba *et al*., 2014; Gupta *et al*., 2010).

Another point which is often not considered in simple analyses, although developed very early by Wells (Wells, 1934), is the evaporation of exhaled microdroplets. Depending on the temperature and relative humidity, microdroplets can rapidly vaporize and have a loss of more than 50 % of their initial size, nearly an order of magnitude in mass. This leads to the formation of very infective “dry nuclei” (Nicas *et al*., 2005) which remain in aerosol form with a much higher viral load than the original droplets. Vaporization is discussed mathematically in the SM.

Based on the work of L. Bourouiba (Bourouiba *et al*., 2014) and others (Drossinos and Stilianakis, 2020) it can be seen than the recommended social distancing of one to two meters is much too low, the puffs emitted by an infected people being able to travel over much larger distances and more particularly to rise due to buoyancy and be sucked into the air intakes of HVAC (Heating Ventilation Air Conditioning) systems, in case of indoor contamination.

Examination of the literature then shows that indoor HVAC systems are prone to homogenize indoor aerosols which justify the use of well-mixed models as in the present paper.

From the biological point of view viruses can be inactivated (*i.e*. lose their infective power) either in aerosol form or on surfaces with a characteristic time (often called the lifetime) which depends strongly of physical parameters such as temperature, humidity or UV radiation field (Ijaz *et al*., 1985; Leclercq *et al*., 2014; van Doremalen *et al*., 2020). Assuming an exponential decrease of virus infectivity with time, introducing it into our calculation is easy, but, based on present knowledge of lifetimes, does not alter the main conclusions on the assessment of the relative risk between outdoors and indoors. Then for the sake of simplicity of the presentation this point is discussed only in the SM.

## 3 Modeling indoor transmission of disease

### 3.1 Exposure level and disease probability

For harmful airborne substances whether chemical (gases), physical (asbestos, soot) or biological (virus, bacteria) it is possible to distinguish between a level of exposure and the probability to develop an illness or even death, (especially in the case of poisonous gases). The level of exposure is often given as a concentration, either in mass or molecules per unit volume, since, multiplied by the pulmonary respiration rate and the time of exposure, it yields a dose which is clearly the risk factor. The probability then of developing a disease (for example cancer from asbestos) must be a strictly increasing function of the dose varying from zero (no exposure) to one (certainty of developing the disease above a given exposure). Most often employment legislation regulates the level of exposure in order to minimize health risk *i.e*. the probability of induced disease.

There have been a large number of attempts to model the transmission of respiratory diseases, most of them being related to indoor situations. The most famous is the Wells-Riley model and its various avatars (Ai and Melikov, 2018; Riley *et al*., 1978; Stephens, 2013) which will be described in the next section. Models use factors identical to the level of exposure as quantum of infection or Minimum Infective Dose and develop links to probability or percentage of infection. The quantum of infection covers the large variety of physical and biological processes involved in infection but for the purposes of the present paper we shall just consider inhalation into the respiratory tract of microdroplets produced by other humans. Thus, in this work the level of exposure to viruses is considered as proportional to the Inhaled Flow Rate of Exhaled Particles, hereafter designed by as IFREP, which is the inhaled flow rate of particles which have already been exhaled by others (including healthy and infected people). We define the Inhaled Dose of Exhaled Particles (IDEP) as the product of IFREP by the time of exposure Δt. Of course, for disease transmission, the proportion of infected people needs to be considered but for comparable situations, the relative level of exposure between indoor and outdoor situations is then the ratio of the respectively calculated IFREP. The time of exposure can readily be considered through IDEP. Note that a similar approach has been developed by other authors for the indoor case only (Issarow *et al*., 2015; Rudnick and Milton, 2003).

By comparable situations, we mean the same population distribution with the same relative number of infected people. The present paper does not compare special indoor environments such as healthcare facilities, especially COVID units, with general outdoor environments. On the other hand, it is perfectly relevant to compare for example, an open outdoor market with a closed indoor supermarket.

Reducing the risk can be achieved clearly by minimizing the level of exposure but knowledge of the probability of infection requires developing a relationship with this exposure level. It will be shown later in this paper that for a Poisson probability law it is easy to link relative probability of infection to the relative level of exposures.

### 3.2 The Wells-Riley model

Following his visionary intuition (Wells, 1934) that respiratory diseases can be due to exhaled airborne particles Wells developed a model of airborne transmission for tuberculosis (and later other respiratory diseases) known as the Wells-Riley model and widely used until now (Ai and Melikov, 2018; Riley *et al*., 1978; Stephens, 2013). Riley was a student and later on a collaborator of Wells. An excellent historical review of their findings and model development can be found in the Master’s thesis of S.P. Johnstone-Robertson (Johnstone-Robertson, 2012). Recognizing that the amount of emitted human aerosols from people known as “infectors” was the equivalent to an exposure level, Wells introduced a quantity that he named the “quantum of infection” proportional to a number of infective airborne particles. The very mechanism by which infective particles trigger a respiratory disease is far from being fully understood, even nowadays. It involves a variety of processes such as deposition of particles in the respiratory track, (Nardell, 2016; Sze To and Chao, 2010). The great advantage of this notion of quantum of infection is that it clearly incorporates this variety of processes without seeking to establish mechanisms. Wells (Riley *et al*., 1978; Wells, 1955) introduced a quantity *q* which is a rate of production of quantum per unit time, per infected person (infector). The equivalent to the dose of exposure, in the notation of Riley 1978, can then be defined as *I* × *q* × *p* × *t*/*Q* where *I, p, t* and *Q* are respectively the number of infectors, pulmonary ventilation rate (volume/unit time), time of exposure and the rate of room ventilation with fresh air. The above quantity was calculated for a stationary state.

As discussed in the previous subsection, exposure and dose level are not the probability to develop a disease. Therefore, Wells and Riley introduced a probability of infection *P* following a Poisson law:

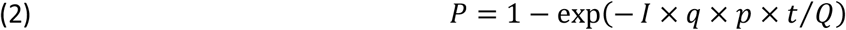

Note that except for the quantum of infection rate *q* and the number of infectors *I*, other quantities in this equation are well known for any disease. Hence, the quantum of infection production rate *q* per infector needs to be determined by epidemiological studies in situations where the number of infectors and infected can be estimated. For new emerging viruses, these quantities are in general unknown at the onset of the epidemic and therefore need to be determined in order to make forecasts regarding the spread of the disease.

### 3.3 A simple homogeneous model of IFREP and IDEP

In this sub-section a simple calculation of the IFREP in an indoor space of volume V (area A, height h) is presented. The situation is depicted in fig. 1.

**Fig 1:**
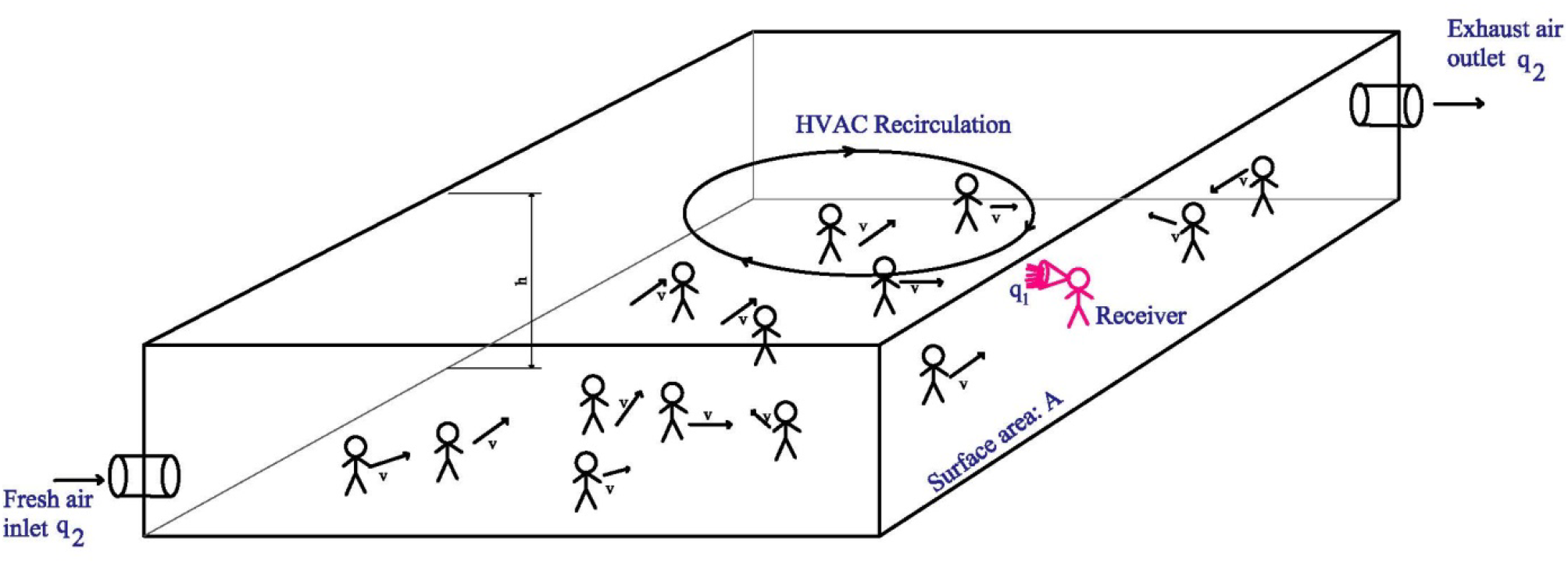
A schematic description of a typical indoor situation.

Let N_p_ be the number of people inside, *N*_*i*_(*t*) the total number of aerosol particles of human respiratory origin inside the volume, it being assumed that recirculation, which is present in most HVAC systems, or by the movements of people, ensure an homogeneous mixing of the particles resulting in a concentration of particles of *n*_*i*_(*t*) = *N*_*i*_(*t*)/*V*. No consideration for the distribution of particle size is done here. For simplicity we shall take this as mono-disperse. The infective power either as “quantum of infection” or MID is not considered either. Again, this is justified by the fact that the purpose of our calculation is just to compare IFREP and IDEP in comparable indoor and outdoor situations. The mean exhaled flow rate of a person is taken as *q*_*1*_ (of course identical to the inhaled rate) and the concentration of particles in this flow will be assumed equal to *n*_*1*_. The flow rate of fresh air introduced in the volume is *q*_*2*_. A typical value for *q*_*2*_ can be taken from HVAC standards (Legg, 2017; Lemaître, 2011) which ideally give the renewal flow rate per individual:

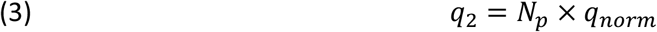

Typical values of *q*_*norm*_ range from 20 to 60 m^3^/h/person.

The mean air exhaust flow rate of the building is equal to the mean inlet flow rate *q*_*2*_ of fresh air considering characteristic time of the problem. As in the Wells-Riley model we do not consider the possible variation of this flow rate with time.

Then the differential equation which governs the temporal evolution of *N*_*i*_ can be written:

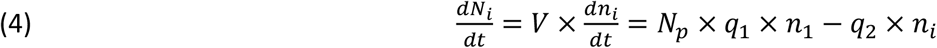

Assuming no exhaled particles at zero time the solution for *n*_*i*_ is straightforward:

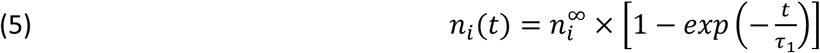

with:

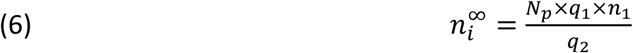

and:

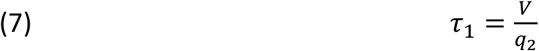

Considering (eq. 2) it is seen that the value of *n*_*i*_ for *t* ≫ *τ*_1_ *i.e*. 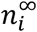 is a function of the norm (inlet fresh air/person) and human respiratory characteristics and not of the number of people:

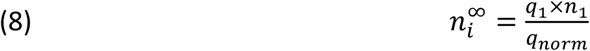

As in the Wells-Riley model this quantity refers to a stationary state. For a person arriving in the building at time *t* ≫ *τ*_1_ the value of IFREP will be 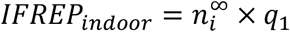 and if he stays there for an interval of time Δ*t* then the value of IDEP will be *IIIP*_*indoor*_ = *IFREP*_*indoor*_ × Δ*t*.

This simple model can be easily extended to the nonstationary case as it will be discussed in the SM. For indoor situation It is quite similar to the so-called “rebreathed air” models (Issarow *et al*., 2015; Rudnick and Milton, 2003).

### 3.4 Inhomogeneous models

The Wells-Riley model is based on two main assumptions: the indoor air that is inhaled by the human receptor is well mixed and at stationary state. The indoor IFREP model developed above is easy to use for the transient state where the concentration of infective particles (or quantum) is increasing as discussed in the SM.

Evaluating the airborne infection risk considering spatial resolution is much more challenging as discussed by Zhang and Lin (Zhang and Lin 2020 and references therein). Some attempt in this way have been made and can involve Computational Fluid Dynamics (Li *et al*., 2018; Vuorinen *et al*., 2020; Zhang and Lin, 2020). However in essence inhomogeneous models are devoted to particular situations, then drawing general conclusions on the relative outdoor versus indoor case seems beyond their possible field of applications.

## 4 Outdoor transmission: A simple airshed model of IFREP

We take now the situation depicted in fig. 1 and remove the walls and ceiling to imagine the same situation transposed “outdoors”. This is depicted in fig. 2. We use here what is called an “airshed concept” which is used in the analysis of city air pollution (Cushman-Roisin, 2012). The problem is analyzed from the perspective of a material balance over a specific part of the atmosphere, the airshed. Although this volume cannot be accurately defined, with the same precision as water in a pool, it is a useful concept. Its definition is of course strongly dependent on atmospheric conditions.

**Fig 2:**
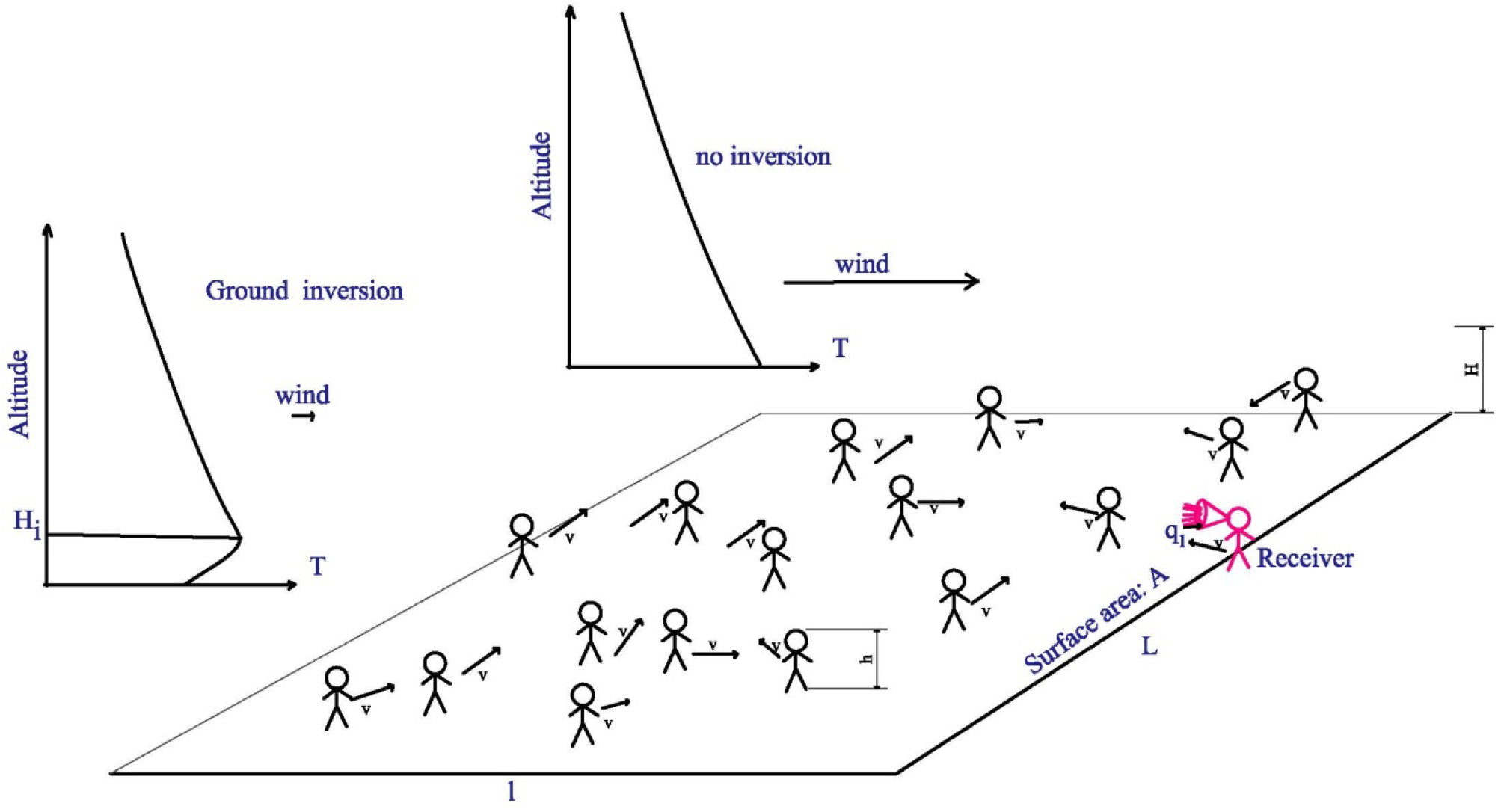
Same situation as in fig. 1 but outdoors with two possible meteorological situations depicted.

The surface of area A has a width along the wind *l* and a length across it, L. The wind velocity is *V*_∞_. Other parameters have the same meaning as in section 3.3 and fig. 1; and *n*_*i*_(*l*) is the concentration of human exhaled particles at the downwind border of the area A.

For typical situations, with *l* comprised between say twenty meters and one kilometer the hydrodynamic time scale *t*_*h*_ = *l*/*V*_∞_ for most usual wind conditions is from a few seconds to a few minutes and mostly well below one hour. Assuming a stationary state and that most of the respiratory human particles are well mixed and will exit the surface area downwind below a typical height *H*, the following conservation equation can be written:

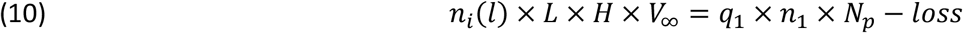

The term “*loss*” takes care of possible particle losses above the height *H* together with lateral ones. Setting this term to zero in fact maximizes the evaluation of the outdoor risk. Then *n*_*i*_(*l*) can be written as:

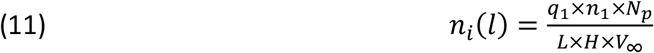

and *IFREP*_*outdoor*_ for and outdoor receiver downwind of the surface A will be: *IFREP*_*outdoor*_ =*n*_*i*_(*l*) × *q*_1_.

Note that minimizing the value of *H* in equation (11) also maximizes directly *IFREP*_*outdoor*_ and therefore the level of exposure which can be a deliberate choice, as discussed below.

Note that eq. 11 can be written as:

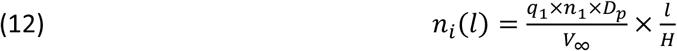

where *I*_*p*_ = *N*_*p*_/(*l* × *L*) is the number density per surface unit of people outdoor.

Determining the quantity *H* = *f*(*l*) is an extremely complicated problem of atmospheric physics. *H* can be defined as a height above which the total flow rate of human exhaled particles become negligible for the airshed balance *i.e*. much lower than its counterpart from ground to this same height. But the choice can also be made to deliberately take a lower value if it leads anyway to a risk much higher than the real one outdoor. Deriving its value from scratch and only basic principles is a virtually impossible task. However, its order of magnitude can be evaluated from a large number of studies in the field of atmospheric pollution, based as well on theory than on experimental observations. Before presenting this evaluation, simple analytical expressions are derived for relative level of exposure and relative probability of infection in the next section.

## 5 Comparison indoor versus outdoor

### 5.1 Simple assessment of relative level of exposure indoor versus outdoor

The purpose of this paper is essentially to assess the relative aerosol contamination risk between outdoor and indoor situation. As stated above, we assume two comparable situations *i.e*. the proportion of infected people is the same with the same characteristics (exhaled flow rate, particle concentration and viral load within the flow) and that “receivers” present the same sensitivity to infection, the relative risk can be evaluated from the quantity that we have defined and evaluated (under simple hypothesis) in the previous sections as IFREP and IDEP. Note that IDEP which is the product of IFREP by a time of exposure Δt is the real factor of risk. Both quantities consider only the continuous inhalation of the bulk air. Crossing (or contact) transmission which can occur when you cross a person but also if you stay “downwind” of this person for a given time can be treated only by inhomogeneous models and is not considered here. This is true for both indoor and outdoor situations.

Then **for the same exposure time**, the relative level of exposure *R* between outdoor and indoor can be estimated as the ratio *IFREP*_*outdoor*_/*IFREP*_*indoor*_ which can then be written as:

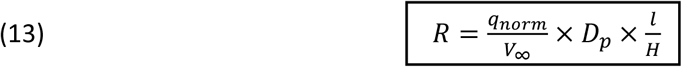

Note that *q*_1_ and *n*_1_ vanish. Under this form the only quantity which refers to the indoor situation is *q*_*norm*_. This is due to the fact that the ventilation rate of fresh air is normally proportional to the number of people indoors. *D*_*p*_ is the number density of people outdoors (person/square meter), *D*_*p*_ = *N*_*p*_/*A*, the quantity *H*/*l* will be discussed at length in the SM where it will be shown that it is linked to the meteorological conditions and to the wind *V*_∞_ itself.

Indeed a correlation between the epidemic and wind as well as atmospheric conditions, (including pollution) which yield the *l*/*H* factor has been observed (Al-Rousan and Al-Najjar, 2020; Rendana, 2020). Correlation is not causality, but the above formula sheds a clear light on the observations.

Note that a different formula can be derived for situations where the indoor ventilation rate does not follow the norm but for the sake of conciseness and clarity this development can be found in the SM.

### 5.2 Probability of infection

As others, including Wells and Riley (Riley *et al*., 1978; Wells, 1955) we use a Poisson law of probability to compare the infection probability indoor and outdoor (respectively *P*_*indoor*_ and *P*_*outdoor*_) for similar situations. (14)

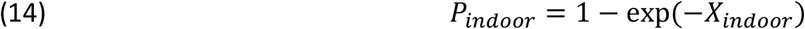

The value of *X*_*indoor*_ in this model has been presented previously and is in fact proportional to IDEP (and therefore IFREP). Clearly it is possible to make a parallel between the quantity that we use in our IFREP calculation and the *X* used in the Wells-Riley model.

The calculations and concepts presented in previous sections allow to calculate a relative level of exposure between outdoors and indoors which can translate directly into a relative value of *X*:

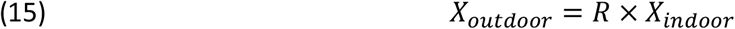

From here, we can derive that, for comparable situation, the outdoor probability of non-infection is linked to the indoor one by:

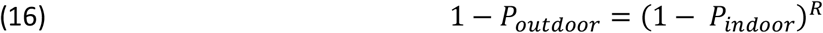

where 1-P is the probability of not being infected. The factor *R*, if ≪ 1, leads to a tremendous advantage to the outdoor in many situations then.

It has to be noticed that for small values of *X*, and therefore of *P*, respective Taylor expansions of the probability show that the ratio of probabilities of being infected reduces to the *R* factor.

### 5.3 Atmospheric conditions, dispersion and choice of ***H*/*l***

The choice of *H*/*l* is clearly central in assessment of relative level of exposure and probability of infection between indoor and outdoor situation. Indoor modeling of a well-mixed homogeneous situation is governed by simple considerations, assuming complete stirring by convection currents in a closed finite volume. Therefore, diffusion does not need to be considered. The outdoor airshed balance is also based on simple considerations but as stated above includes an unknown quantity *i.e*. the height of the airshed. The determination, even approximative, of its value obeys different phenomena, generally much more complicated and harder to consider than in the indoor situation. First the advection by the wind, which is always present, even be it very weak and not constant, in intensity as in direction. Besides this transport by advection, atmospheric turbulence contributes to the dispersion by so-called eddy diffusion (much more efficient than molecular diffusion). This important point is related to the stability of the atmosphere (see SM), which in turn depends on meteorological conditions, solar exposure, and thermal radiation from the ground and from the clouds.

*H* could have been taken as the so-called mixing height (Holzworth, 1974) which can be defined as the vertical height over which an unstable parcel of air taking off from the ground will rise (see below). Therefore, it corresponds in fact to the maximum height of the atmosphere where mixing of pollutant occurs. This quantity can change from zero in situations of inversion to thousands of meters (Holzworth, 1974). Therefore, it seems more reasonable for the present case to use a model of the dispersion of gaseous pollutants on a more reasonable length scale.

There is a large literature on the plumes and puffs emitted from smoke stacks and the estimation of pollutant concentration downwind due to dispersion, see for example the excellent course (Pilat, 2009) and the review by Holmes and Morawska (Holmes and Morawska, 2006). There are also numerous papers linked to biology which treat of odor dispersion, for example of insect pheromone downwind (Farrell *et al*., 2002). Our problem can be thought of as a field with several analogs to a small smokestack producing airborne particles, the human emitters. We make use of what is known of the vertical length of dispersion, extremely dependent of meteorological conditions, to evaluate *H*. In the sole purpose of lightening the text of the present article we refer the reader to the SM for further reading on atmospheric conditions and stability and their influence on the dispersion of plumes and puffs. In this document and following Pasquill (Pasquill, 1961) classes of atmospheric stability are defined ranging from A (very unstable) to F (very stable), D referring to a neutral atmosphere (as defined in the SM). In the case of a Gaussian vertical dispersion it is shown that for typical lengths involved in many practical situations (outdoor market for example) *H* is proportional to *l* which implies the choice of a constant value for *H*/*l* shown in table 1.

**Table 1:** adopted values of *H*/*l* for vertical Gaussian dispersion and various meteorological conditions (see SM)

|  | A | B | C | D | E | F |
| --- | --- | --- | --- | --- | --- | --- |
| $H/l$ | 0.2 | 0.097 | 0.074 | 0.051 | 0.034 | 0.023 |

However not all the plumes can be considered as being described by a vertical Gaussian dispersion, in some meteorological situations there are hardly any vertical dispersions (situation corresponding to “fanning” and “fumigation”, described in SM). In this very special case, which corresponds to a very stable and strong inversion situation, we assume a constant value for *H* that will be taken as the mean height of an emission source *i.e*. 1.5 meter.

### 5.4 Numerical application and discussion

In the present section we assume that the indoor ventilation rate is fixed accordingly to the norm. In table 2, some values of factor *R* obtained for a few typical meteorological conditions and outdoor person density are listed for the case of a vertical Gaussian dispersion.

**Table 2:**
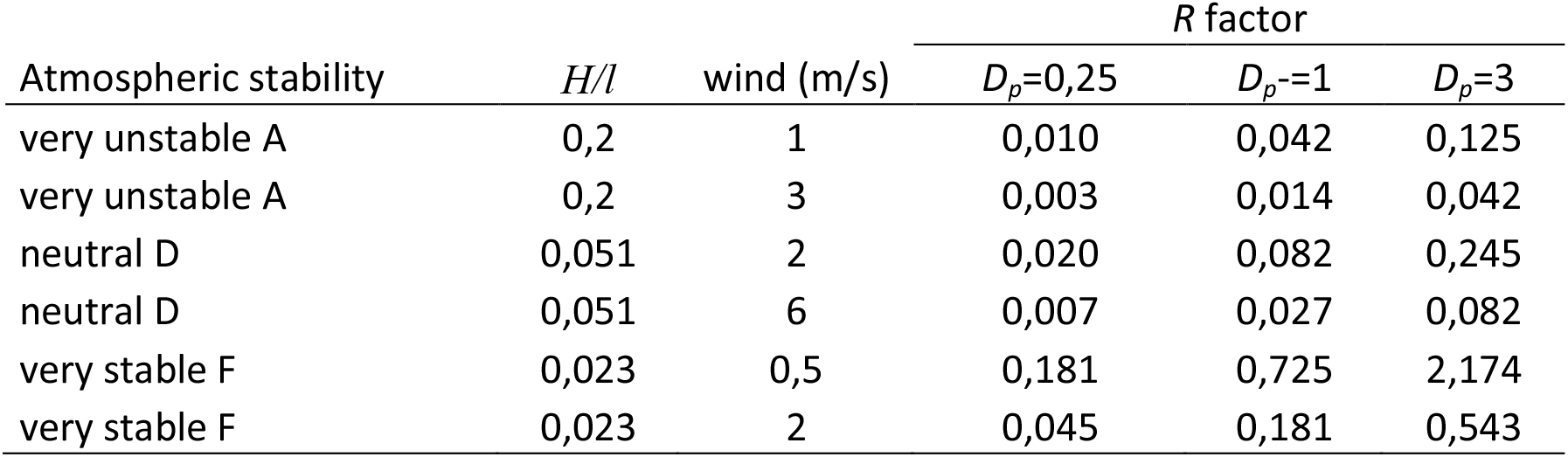
relative risk factor *R* (outdoor/indoor) for Gaussian vertical dispersion, various atmospheric conditions and people density.

| Atmospheric stability | $H/l$ | wind (m/s) | $R$ factor | | |
| --- | --- | --- | --- | --- | --- |
| | | | $D_p=0,25$ | $D_p=1$ | $D_p=3$ |
| very unstable A | 0,2 | 1 | 0,010 | 0,042 | 0,125 |
| very unstable A | 0,2 | 3 | 0,003 | 0,014 | 0,042 |
| neutral D | 0,051 | 2 | 0,020 | 0,082 | 0,245 |
| neutral D | 0,051 | 6 | 0,007 | 0,027 | 0,082 |
| very stable F | 0,023 | 0,5 | 0,181 | 0,725 | 2,174 |
| very stable F | 0,023 | 2 | 0,045 | 0,181 | 0,543 |

The first one is the situation of very unstable atmosphere which corresponds to heated ground and can be found in the daytime in summer at mid-latitudes or in the dry season in a tropical area. Another situation is a stable but moderately windy condition, corresponding for example to a winter day at mid-latitudes. The last one corresponds to a very stable atmosphere with low wind. In this table, we have taken a mean value of *q*_*norm*_ of 30 cubic meter/hour/person. Note that the wind here is taken as its mean value at 10 meters of altitude. In the real world, the wind has a vertical profile, starting from zero at the ground and increasing with altitude (Wikipedia, 2020), see also the discussion in the SM. Of course it is also a temporal mean (on ten minutes) as the wind is always turbulent and variable.

In table 3, results are presented for a constant height of dispersion (1.5 meters) versus downwind distance to emitters. This corresponds to special and very stable meteorological conditions as discussed in the previous section.

**Table 3:** relative risk factor *R* (outdoor/indoor) in strong inversion and low wind conditions for various lengths along the wind *l* and people density.

| wind (m/s) | $l$ (m) | $R$ factor | | |
| --- | --- | --- | --- | --- |
| | | $D_p=0,25$ | $D_p=1$ | $D_p=3$ |
| 0,5 | 50 | 0,14 | 0,56 | 1,67 |
| 0,5 | 100 | 0,28 | 1,11 | 3,33 |
| 2 | 50 | 0,03 | 0,14 | 0,42 |
| 2 | 100 | 0,07 | 0,28 | 0,83 |
| 2 | 200 | 0,14 | 0,56 | 1,67 |

The main conclusion that can be drawn from tables 2 and 3 is that, most generally, outdoor risk is much less than indoor (eventually by orders of magnitude). Only situations of inversion with low wind and a very stable atmosphere, which are prone to atmospheric pollution, could promote an outdoor transmission close to that indoors, especially in crowded area. If this situation occurs the “fresh” air that is introduced from outside in buildings could be already “polluted” leading to a strongly enhanced indoor airborne transmission of the disease. In the authors opinion this could explain some observed outbreak of the epidemic depending on geographical area and meteorology as shown in a recent paper (Rohrer *et al*., 2020). Note the importance of the wind factor and of the outdoor people density.

## 6 Some markers of risk outside and inside

### 6.1 anthropogenic aerosols outdoors

It has been observed especially in Italy (Rohrer *et al*., 2020; Zoran *et al*., 2020) a correlation between pollution by particulate matter and outbreak of the epidemic. It has been suggested that in a synergistic effect pollution can increase the infective power of virus by agglomeration of airborne infective particles with PM particles. It is interesting to note that after the discovery by Robert Koch in 1882 of the mycobacterium responsible of tuberculosis it was first thought that tuberculosis was spread by the breathing in of dust particles contaminated with dried mycobacterium tuberculosis laden sputum (Johnstone-Robertson, 2012) an idea which revealed to be wrong. As discussed by Doussin (Doussin, 2020) and in the SM the typical characteristic time of agglomeration in the atmosphere is far too long to allow such an agglomeration within the atmosphere. It is known that bacteria and viruses can be found in atmospheric aerosol (Kalisa *et al*., 2019), but most of natural aerosol are formed by ground surface abrasion by the wind which explain the observation. If the ground is contaminated, then it could of course be the case for particles formed by abrasion.

Correlation is not causality but the observation of peak of pollution due to particulate matter is clearly linked with special meteorological conditions prone to enhance this pollution. Following the present work, we suggest that the cause of **the outbreak of epidemics is these meteorological conditions and not the particulate matter pollution itself**. As meteorology is predictable a few days in advance this could be used for public recommendation and alert.

### 6.2 Carbon dioxide indoor

In air exhaled by human, carbon dioxide has a much larger concentration (4-5 %) than in fresh outdoor air (section 2.1). Therefore several authors (Rudnick and Milton, 2003) have proposed to monitor CO_2_ levels in indoor situation as an indicator of the risk of infection. In fact the simple model developed for IFREP in section 3.3 can be readily applied to indoor CO_2_. Noting [*CO*_2_] the concentration of CO_2_ in ambient air it leads to:

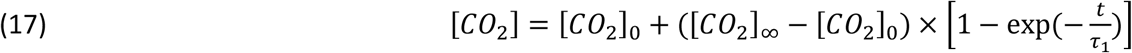

where [*CO*_2_]_0_ is the initial concentration of C0_2_ in fresh air and [*CO*_2_]_∞_ the concentration at stationary state which writes:

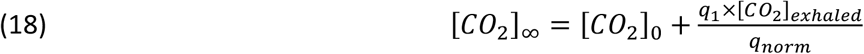

where [*CO*_2_]_*exhaled*_ is the concentration of CO_2_ in the exhaled air.

The characteristic time is the same than for IFREP hence the interest of monitoring CO_2_. With typical value of respiratory parameters and *q*_*norm*_ it is found that at stationary state the amount of CO_2_ in air is easily twice the fresh air one, making monitoring easy. An increase of *q*_*norm*_ by an order of magnitude yields only an increase of around 10% of the concentration at stationary state, therefore monitoring would be still possible with precise sensors.

## 7 Conclusions and recommendations

The last months have seen an extraordinary inflation of papers dealing with the Covid-19 and its transmission. A great amount of these papers deals with correlation and observation and not with quantitative models of the physical processes of transmission. The fact that correlations are not causalities makes difficult their use for public decisions to mitigate virus spread preserving as much as possible social life and the economy. For example, it is clear that a measure to reduce pollution by particulate matter emitted by combustion (as taken for transportation in urban area) will have no effect on the spreading of the disease in view of the arguments developed in section 6.1 and in the SM.

Airborne transmission of COVID-19 is now widely recognized, and this has led public authorities to recommend or impose the wearing of mask in the general population, certainly an excellent mitigation measure as shown by observation. However, when it comes to the question to know when to wear it the answer is far from being so evident, although it is clear that wearing it night and day and in any circumstances is not realistic

In this paper, funded on quite simple calculations, we have presented a quantitative assessment of the relative risk of airborne virus transmission for the outdoor versus indoor situations. Calculations result in remarkably simple formulas which, considering the science of atmospheric physics and pollution allows to assess the relative risk between indoor and outdoor situation. The simplicity of this derivation could be criticized, and its application is most often based on proportionality rule. On another hand, it makes the beauty of these simple formulas which, through each of the parameters involved, makes sense.

First and, as discussed in section 5.3, it shows that even in crowded area the outdoor risk is much less than indoor. From this point of view, it has to be noted that some decisions taken by public authorities could have appeared as absurd to the man in the street. This opinion just based on common sense is confirmed by the present study. Examples are very numerous ranging from the lockdown of open markets when indoor supermarkets were open to the public, or prohibition of a variety of outdoor sport and exercise. Let us point out that such lockdown and prohibition can have profound impact on health of citizen and economy. Note that in the present paper we have not considered the phenomena of droplet evaporation indoor which is strongly enhanced by the low relative humidity due to heating in winter time, and clearly will lead an even higher indoor risk, reinforcing our conclusion.

Considering mitigation measures it is clear that wearing mask, especially indoor, is an important way to reduce risk of contamination (as well contact than airborne). However, and as noted by Morawska *et al*. (Morawska *et al*., 2020) other measures should include strongly increasing ventilation which means increasing the norm of fresh air renewal per person by an order of magnitude either in HVAC systems or by natural ventilation (Escombe *et al*., 2007). If this is not possible, apparatuses allowing to sterilize indoor air should be envisaged, however, to be efficient, here again they would need to be able to treat a flow rate at least an order of magnitude higher than the present norm.

Following our study, we are led to believe that the fact that Africa, especially sub-Saharan, has not been stricken by the disease so much than rich mid-latitude country, is linked to climatic factors (very unstable atmosphere) together with an outdoor way of life (for example outdoor market instead of climatized indoor supermarket). Due to the low gross national product per habitant the use of climatized air is also much less common than in richer countries of same latitude. On the contrary a situation of temperature inversion can occur within the day, especially in wintertime for mid latitudes where the sun is low in the sky and supplies less warmth to the Earth’s surface. Either radiation or the so-called subsidence inversion (*i.e*. above the ground) can inhibit vertical dispersion and act as a lid and trap cold air at the ground. These effects can be strongly amplified in mountain valleys. We suggest that, in these geographical areas prone to pollution by secondary PM 2.5 which include large urban areas with collective housing and towers, monitoring this pollution, together with meteorological forecast, could be a way to alert the population of risky days and to reinforce mitigation measures for short period of time. Such suggestion has already been done for indoor CO_2_ level (Rudnick and Milton, 2003).

To finish we want to emphasize that the spreading of the disease is an extraordinarily complex phenomena certainly not restricted to the airborne transmission way, although this way can make the difference for the effective reproduction number leading or not to an epidemic burst. The modest contribution of the present paper is an attempt to quantitatively assess the relative risk linked to aerosol between outdoor and indoor situation. We hope that it will encourage atmospheric physicists and pollution experts to tackle the outdoor dispersion problem of respiratory produced aerosols in more details.

## Data Availability

no additional data available

## Acknowledgements

The authors wish to acknowledge Allan Crouvizier, Alexandre Modesto and Alexei Potapov for helpful discussions and bibliographic watch.

## Supplementary materials

### S1. Micro droplet evaporation

An aerosol is released while coughing, sneezing, speaking or simply breathing. The corresponding droplets, mainly in the range from a fraction of micrometre to one hundred, enclose the viral content. It might be believed that most of the mass of these droplets fall to the ground and therefore that there is no airborne contamination. As already recognized by Wells in 1934 (Wells, 1934) this is false because these water droplets can evaporate in the ambient atmosphere; they shrink until their falling speed becomes so small that they are carried by the small movement of the atmosphere, would it be very quiet.

Quantitatively, due to the Stokes law, the droplets of radius *r* reach a limit speed in the (viscous) air of

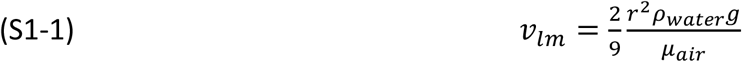

where *ρ*_*water*_, *g* and *μ*_*air*_ are respectively the volume mass of water, gravity acceleration and air viscosity. This means that a non-evaporating droplet of diameter 100 μm falls from a height of 1.5m in 5 s; with a diameter of 30 μm, 55 s are necessary; and with a diameter of 10 μm, the falling speed lowers to 3 mm/s so that this droplet may be dragged by the movements of the atmosphere.

Now consider the evaporation. The models are quite complex, and we refer for instance to reference (Lefevre and McDonel, 2017) for the details. The main result is that the squared diameter of a drop shrinks linearly in time:

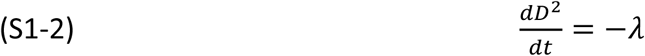

where *λ* depends in particular on the relative humidity *RH* and is falling to 0 in a saturated air. Hence plugging in the limiting fall speed above, one finds:

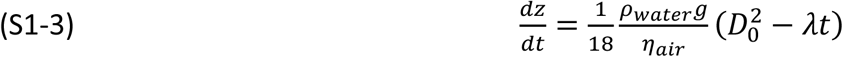

Integrating this differential equation up to the life time of the droplet 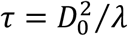, one finds the falling height *h* between its production and its complete drying

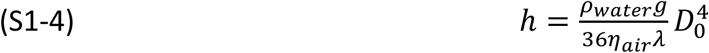

If this height is greater than the production height (typically 1.5 m), the particle deposits on the ground; otherwise, it becomes so light due to the evaporation that it participates to airborne contamination.

We do not enter in the evaluation of the parameter *λ* which expresses an equilibrium between heat transfer towards the droplet and water vapour mass transfer from the droplet. It depends on the temperature which governs the saturated water vapour pressure near the droplet surface and on *RH* which commands the mass transfer of water vapour from the droplet (see (Lefevre and McDonel, 2017) for the detailed formulae). Hence we give only a final table (T1-1) below for the results at the temperature of 20 °C for various *RH*. The second column displays the limiting diameter under which the droplet evaporates completely before reaching the ground supposed to be 1.5 m under the droplet production. Below this diameter, all droplets would participate to airborne transmission; the airborne dispersed droplet proportion that would be obtained with these calculation hypotheses is given in the last column, assuming a flat distribution of droplets between 30 μm and 100 μm. The interest of table T1-1 is to show the importance of relative humidity on droplet evaporation.

**Table T1-1.**
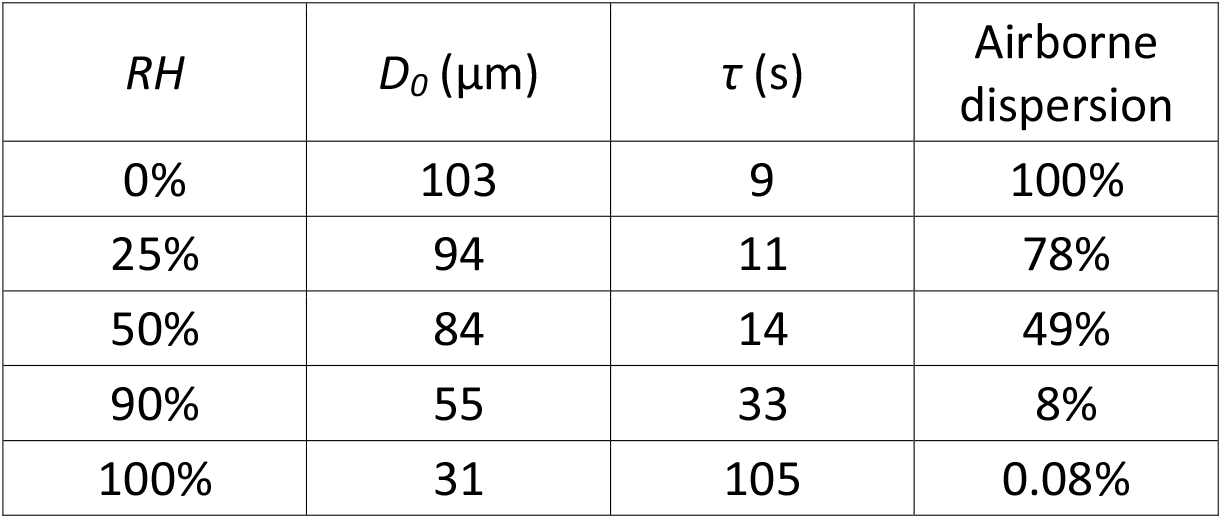
Droplet airborne evaporation for pure water.

| $RH$ | $D_0$ ($\mu m$) | $\tau$ (s) | Airborne dispersion |
| --- | --- | --- | --- |
| 0% | 103 | 9 | 100% |
| 25% | 94 | 11 | 78% |
| 50% | 84 | 14 | 49% |
| 90% | 55 | 33 | 8% |
| 100% | 31 | 105 | 0.08% |

Note that indoor conditions lead generally to dry air (*RH =* 15-20%), enhancing airborne dispersion while wet outdoor conditions favour deposition. There are a very large number of papers in the scientific literature on this topic, see for example (Morawska, 2006).

The above results assume that the non-volatile part of the droplet can be neglected which is not true. Considering this point, it can be shown that the final diameter cannot shrink below 26- 44% of its initial value, see for example (Nicas *et al.*, 2005), which implies that for most of the diameter in table T1-1 the dry nuclei will finally fall to the ground. This final diameter corresponds to “dry nuclei” with a complicated composition (salt, protein, viruses). Note that there is no reason to assume that these dry nuclei are hard solid spheres. It is known that evaporation of solutions can lead to hollow or fractal structures which will have a much higher drag coefficient than a compact sphere, leading to an easier aerosolization.

### S2- Transient effects and time of inactivation for the indoor case

The following equation, developed in the main paper, determines the indoor temporal evolution of the concentration of inhaled particles *n*_*i*_ that have been exhaled by human emitters:

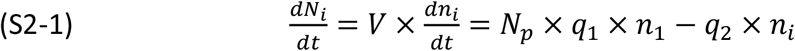

where V is the building volume, *N*_*p*_ the number of people, *q*_1_ the mean exhaled flow rate per person, *n*_1_ the concentration of particle in the exhaled air.

Its solution takes clearly into account transient effects which occur when the time t is not ≫ *τ*_1_ with *τ*_1_ defined by equation (S2-3). This is the case for example at the opening of a supermarket. Of course, as the flow rate of fresh air *q*_2_ could be regulated as a function of the number of people following equation (S2-1) this could make transient analysis rather complicated. However, in many cases this flow rate is adjusted constant, taking into account a “normal” number of people. Equation (S2-1) yields for *n*_*i*_:

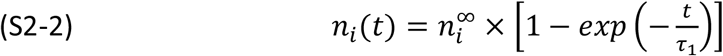

and involves mainly:

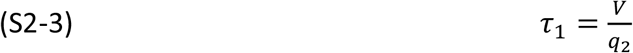

and:

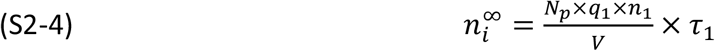

Considering some typical values leads to typical times *τ*_1_ around one hour or below. Clearly the risk is higher toward the end of the day. Note that monitoring CO_2_ is clearly a good way to track transient effects. These transient effects have already been studied in the literature (Rudnick and Milton, 2003).

Another point which could introduce indoor time dependent effect is the time of inactivation (or lifetime) of a virus. If we assume that infective particles loss their infective power with an exponential lifetime *τ*_2_ then it can be added a term in equation (S2-1).

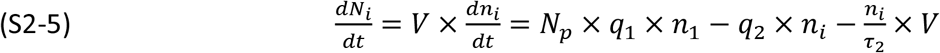

Of course, equation (S2-5) considers all particles of human origin, infective or not. But if we consider that infective particle concentration is just proportional to the total particle concentration, and infector number proportional to the number of people *N*_*p*_, with the same coefficient of proportionality, then it is justified to consider that equation (S2-5) mirror the equation for infective particles and that the derived *n*_*i*_ can be used to calculate a modified IFREP (Inhaled Flow Rate of Exhaled Particles) which takes into account the lifetime. Doing this, results in a modified characteristic time *τ*_*l*_ and stationary concentration value 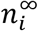.

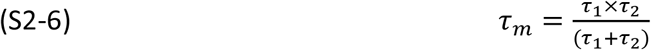

and:

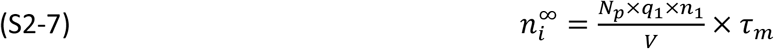

Note that *τ*_1_ can also be written:

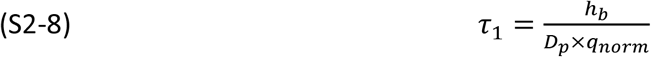

where *h*_*b*_ is the ceiling height of the building, *D*_*p*_ being as in the main paper the people density.

In many situations this leads to *τ*_1_ ≪ *τ*_2_ hence *τ*_*l*_ ∼ *τ*_1_ which means that there is little influence of the virus inactivation on the stationary concentration of particles to consider for the comparison of outdoor to indoor situation, the principal purpose of the main paper.

Note that outdoor for most situation the hydrodynamic time *τ*_*h*_ = *l*/*V*_∞_ is such that viruses have no time at all to be inactivated (*τ*_*h*_ ≪ *τ*_2_).

### S3- Evaluation of the relative level of exposure for any indoor ventilation rate

In the main paper we have established a simple formula for the relative level of exposure between outdoor and indoor situations:

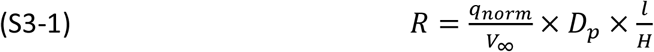

Under this form the only quantity which refers to the indoor situation is *q*_*norm*_. This is due to the fact that the ventilation rate of fresh air is normally proportional to the number of people indoors. *D*_*p*_ is the number density of people outdoors (person/square meter), *D*_*p*_(*outdoor*) = *N*_*p*_(*outdoor*)/*A*, the quantity *H*/*l* is the ratio of the height of dispersion to the length along the wind in the area *A*.

Note that if the indoor ventilation rate is not fixed accordingly to a number of persons then *R* can write:

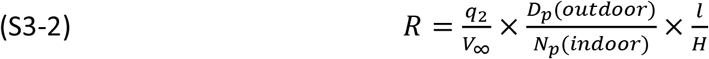

Since *q*_2_, the building fresh air renewal is (as stated above) *q*_2_ = *q*_*norm*_ × *N*_*p*_ Note that *R* can also be written:

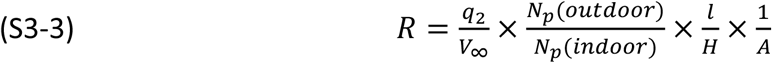

Again in both equations we can see that the parameter difficult to evaluate for the relative level of exposure is the height of dispersion *H*, completely dependent of meteorological and climatic conditions.

### S4 Correlations between “increase of contamination” and “fine particle pollution episodes”

A correlation between episodes of fine particle (PM hereafter) pollution and the increase in contamination cases has been observed on several occasions (Rohrer *et al*., 2020; Zoran *et al*., 2020). It could be deduced from this that atmospheric PM pollutants act as vectors for viruses because of surface aggregation and reaction phenomena. However, it can also be stated: **“Atmospheric conditions favourable for keeping pollution, such as soot particles in the air are also favourable for keeping viral particles in suspension**”.

Therefore, even in an un-polluted zone, these same atmospheric conditions favour the transmission of the virus.

Of course, atmospheric pollution is not healthy for the lungs of individuals and could possibly predispose them to suffer more to viral contamination.

However, the calculation presented below shows that it is indeed the second statement, *atmospheric conditions favourable to aerosols*, that should probably be retained. Thus, the message concerning airborne transmission of the virus should not be mixed in with that on pollution: in a contaminated area with specific meteorology and geography, airborne transmission could be effective outdoor with or without PM pollution.

#### The calculation

This calculation has been performed considering a “gas of particles” where between two interactions, the particles follow a straight trajectory. In reality, particles undergo “Brownian” (or diffusion) motion where the trajectories are not straight, however, taking this complexity into account changes the results by less than an order of magnitude. The calculation is performed with the following data:

- For the sake of the calculation, pollution particles are considered to have a density *ρ* of 2000 kg/m3 (this is typical soot density but could be another material).
- Their diameter will be taken equal to three successive values i.e. 10, 2 and 0.2 *μ*m.
- The weight of pollutants is 40 *μ*g/m^3^
- The air temperature T = 290K (17°C).

It should be noted that this weight of pollutants per unit volume is what is retained in the standards (not the number of particles/m^3^) and what is usually measured. Obviously, the choice of a size of the particles will then considerably influence their calculated concentration per unit volume. Usually standard by weight is given per size of particles, i.e. the allowable total weight of particles below a certain size. Here we use the maximum weight concentration of PM10 as given by the European directive 2008/50/CE.

From the above data, supposing that the particles are mono-disperse and in the assumption of a “gas of particles” the number of particles per unit volume n (particles/m^3^) can be calculated together with the following quantities:

- Their cross section σ=πd^2^/4 (m^2^)
- The weight of a particle (m=4*ρ*πr^3^/3, r =d/2)
- A thermal agitation speed 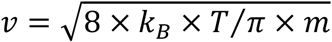 with *k*_*B*_, Boltzmann’s constant, m particle mass
- The agglomeration reaction rate coefficient in m^3^/s, *i.e. k*_*agg*_ = *σ* × *v*
- The characteristic reaction time for agglomeration (*τ* = 2/*k*_*agg*_ × *n*) seconds)

#### From all these parameters, the most significant is the characteristic time *τ*

The presence of air will change the results somewhat but not by an order of magnitude. The input parameter that affects the results most is the size retained for the particles (through the derived number concentration from the adopted mass concentration).

The calculation of *k*_*agg*_ and hence *τ* is an order of magnitude in agreement with more sophisticated calculations that can be found in the literature, taking into account the influence of the air, Brownian motion, Van der Waals forces and the Knudsen number. Such a refined calculation is shown in figure F-S4-1.

**Figure F-S4-1:**
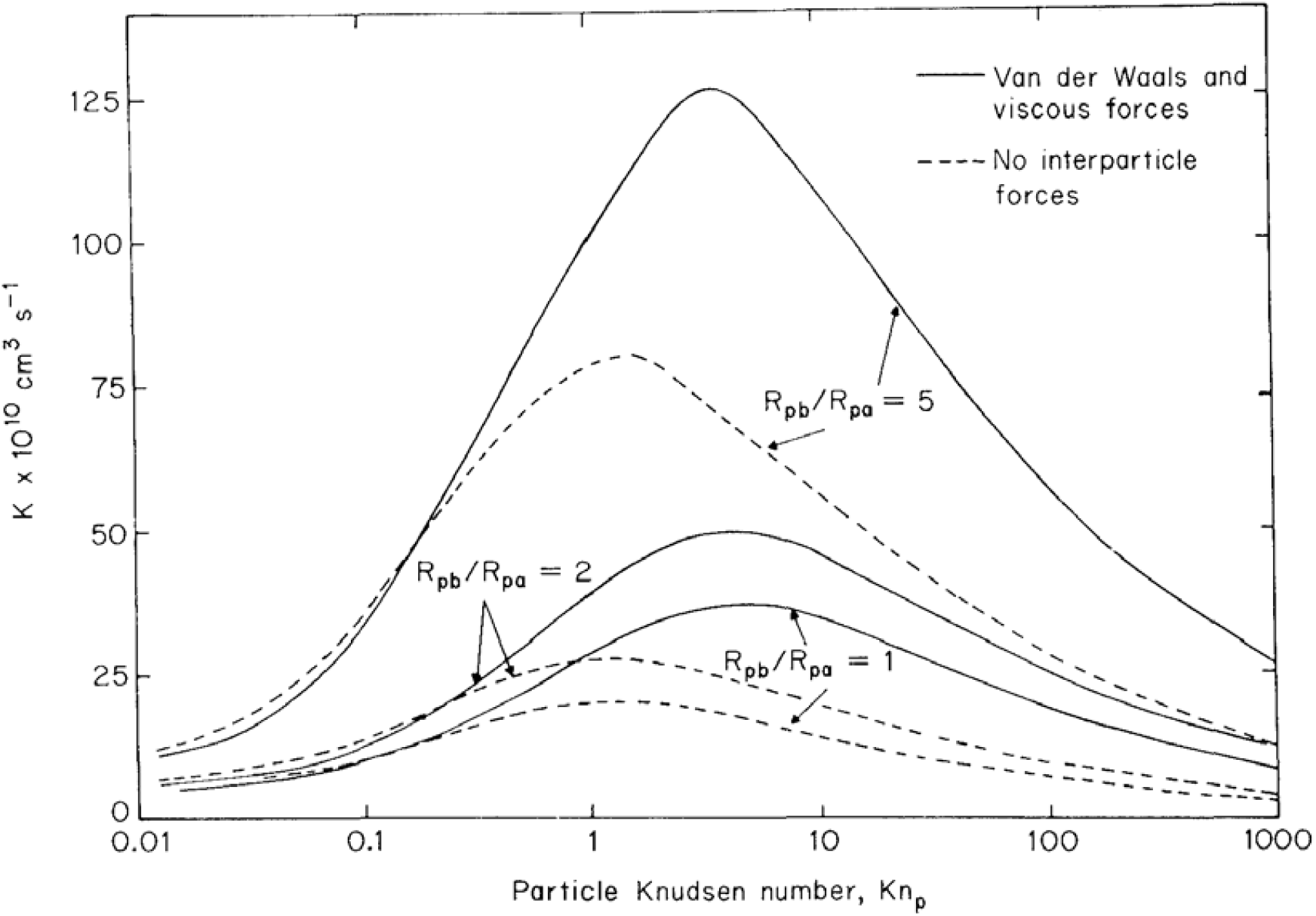
Variation of the agglomeration coefficient as a function of the Knudsen number for different assumptions (from reference (Fuchs, 1989).

Thus, with the input parameters given above, the characteristic time for coagulation is very long. If the size of the given particles (e.g., soot) is 10 microns, it is on the order of a century, for 2 microns, on the order of one year and for 0.2 microns (200 nm), about 2 days. This must be compared with the lifetime of a virus in an aerosol particle (dry or wet nuclei).

These order of magnitude calculations are for a mono-disperse (single size) aerosol and a concentration equal to the maximum weight concentration of PM10 as given by the European directive 2008/50/CE. It should be noted that exceeding reasonably this standard value does not change the conclusion, which is in agreement with the discussion made by other researchers (Doussin, 2020).

### S5- Atmospheric considerations

#### The wind

The meteorological wind is normally measured ten meters above ground surface. Usually, its horizontal component is much higher than the vertical one. Close to the ground its variation follows a logarithmic law which includes a variety of parameters such as the roughness length z_0,_ the friction velocity 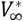 and displacement height D. z_0_ depends on the nature of the surface and is usually much lower than ten meters for open lands with few barriers such as trees or buildings. In this case the wind around 2 m height is quite close (within a factor of 2) to the 10 m wind. D is grossly a displacement of the ground surface, for example in the case of a forest.

It is beyond the scope of the main paper and of this supplementary material to examine the variety of situations that can be found in real circumstances. Therefore, wind values refer always to the meteorological ones and calculations does not consider the complexity linked to tall buildings for example.

#### Atmospheric stability

Vertical dispersion is strongly due to convective instabilities induced by the thermal profile of the atmosphere. Let us recall the argument. If a volume of air at altitude *z* is displaced adiabatically by an amount *δ*_Z_, a hydrostatic change of pressure is induced with altitude. This yields a change of temperature with altitude which can be evaluated by thermodynamics to be *δT* = −Γ × dz with:

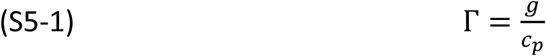

Where g is the acceleration of gravity and *c*_*p*_ the specific heat of air at constant pressure. The quantity Γ is known as the dry adiabatic lapse rate. If this change of temperature *δT* is less than the real decrease of temperature in the atmosphere, the displaced volume is denser and will return by buoyancy to its original place: the atmosphere is stable. Otherwise, the displacement will be amplified: there is a lot of up and down movements tending to restore the mechanical stability. In this situation of instability, pollutants are also moved up and down, amplifying the length of dispersion *σ*_*z*_.

**Figure F-S5-1:**
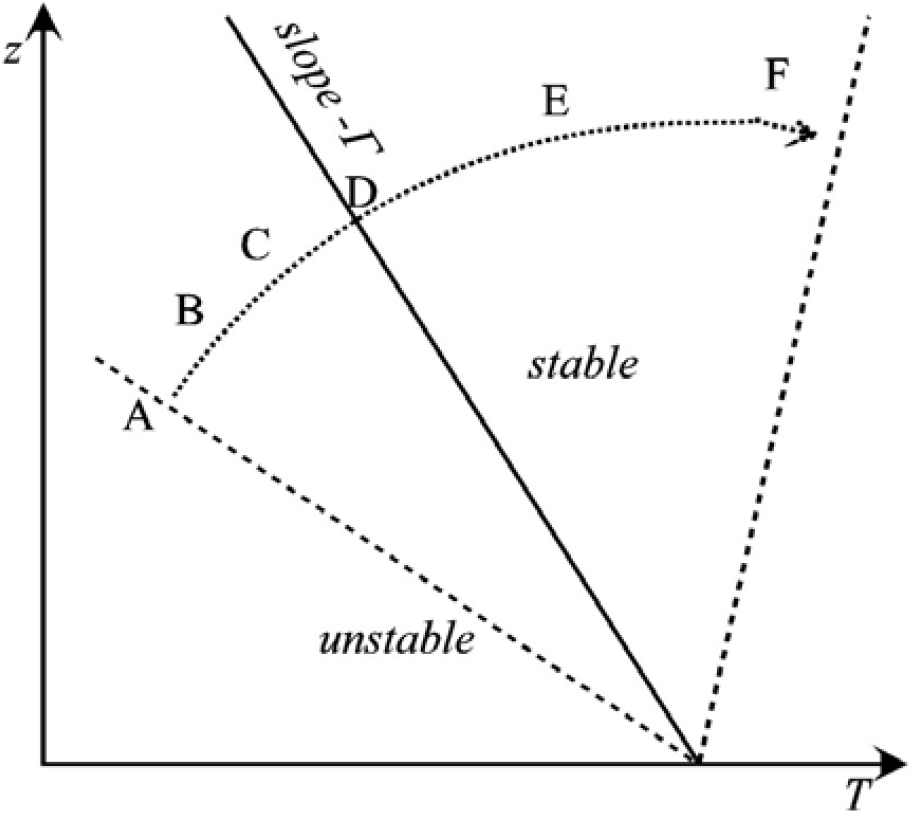
atmospheric stability following the temperature lapse rate. D is the neutral case.

**Table T5-1:**
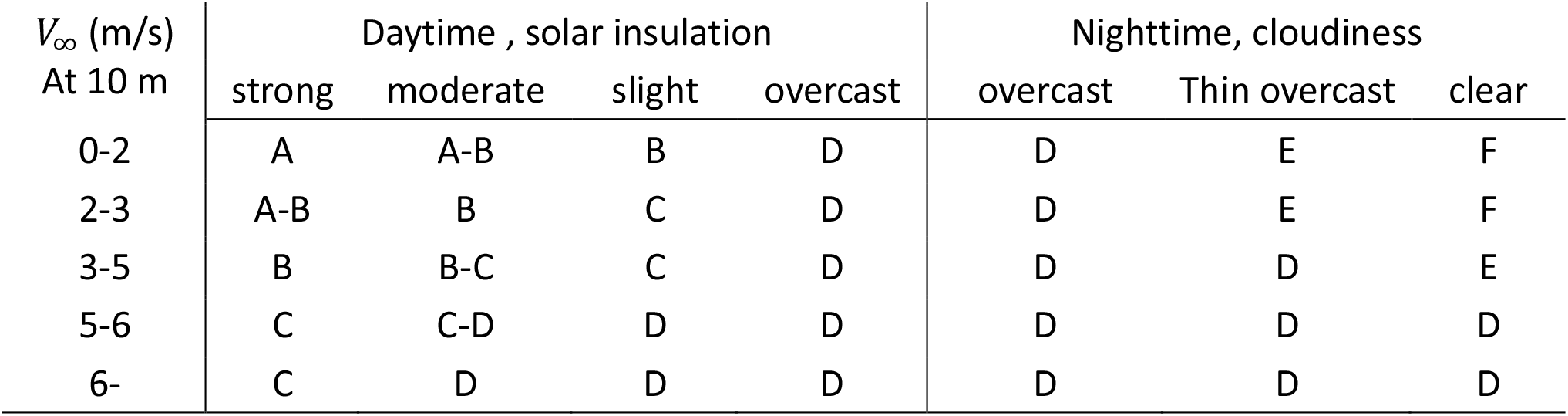
The Pasquil-Giffort-Turner class of atmospheric stability following meteorological conditions. Neutral (D) conditions prevail nights and days for overcast conditions.

| $V_{\infty}$ (m/s)<br>At 10 m | Daytime , solar insulation | | | | Nighttime, cloudiness | | |
| --- | --- | --- | --- | --- | --- | --- | --- |
|  | strong | moderate | slight | overcast | overcast | Thin overcast | clear |
| 0-2 | A | A-B | B | D | D | E | F |
| 2-3 | A-B | B | C | D | D | E | F |
| 3-5 | B | B-C | C | D | D | D | E |
| 5-6 | C | C-D | D | D | D | D | D |
| 6- | C | D | D | D | D | D | D |

Common atmospheric conditions have been classified (the so-called Pasquil– Gifford– Turner scheme, (Pasquill, 1961; Gifford, 1961; Turner, 1994) from A to F, running from very unstable conditions to very stable ones; D is the neutral case *δT* = −Γ × dz. They are shown in table T-5-1 and correspond to figure F-S5-1.

Explanations of this table are rather simple. During the day, sun exposure will heat up the ground, leading to higher temperatures near it and therefore a strong decrease of temperature with altitude; hence *dT*/*dz* ≪ −Γ with *dT*/*dz* < 0 leading to high instability, *i.e. a* large characteristic length of dispersion. Conversely, in a clear night, heat is released by the ground without being trapped by cloud cover, leading to high stability. The influence of the wind speed *V*_∞_ is explained by stirring the atmosphere which is rapidly led to recover the equilibrium thermal profile. Note that a situation of inversion can occur within the day, especially in wintertime for mid latitudes where the sun is low in the sky and supplies less warmth to the Earth’s surface. A so-called subsidence inversion, *i.e*. above the ground, can act as a lid and trap cold air at the ground. These effects can be strongly amplified in mountain valleys.

#### Dispersion of pollutants downwind of a stack

Downwind of a stack there is a stream of polluted air in the form either of puffs or of a plume. Plumes are classified in a variety of ways, depending on meteorological conditions, as shown in figure F-S5-2.

Assuming a quantitatively stochastic random walk leads to a Gaussian distribution downstream of a stack which has been used widely in the field and is appropriate for a number of situations. Description of spatial concentration of pollutants takes the origin at the bottom of the stack, x the distance downwind of the stack, y and z respectively lateral and vertical coordinates. Lateral and vertical dispersion lengths *σ*_*y*_ and *σ*_*z*_ are introduced. They are different since they are produced by different causes. With several sources, lateral dispersion helps to mix and homogenize the particle concentration and is not important for our purpose. We are interested in the vertical dispersion which is strongly affected by the stability of the atmosphere which has been discussed previously.

Assuming a height *z*_*c*_ for the center of the pollution cloud, the polluting concentration at a distance *x* from the source can be expressed as (Turner, 1994):

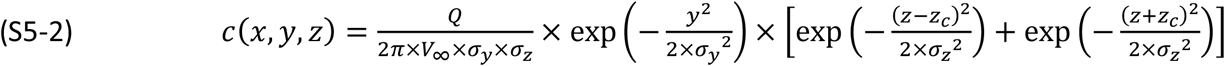

Now the problem is to evaluate *σ*_*y*_,, *σ*_*z*_ as function of *x*, and especially *σ*_*z*_ which is the major parameter for an evaluation of *H. z*_*c*_ is the center of the plume and Q the source term in unit (particles, mass or others) per second consistent with the concentration unit.

The vertical dispersion will depend strongly on the conditions. In the neutral case D, pollutants are emitted in a cone (“coning” in the above figure), due to similarity considerations. When the stability increases (F), the cone angle shrinks, showing better stability (parcels of deviant air are more strongly pushed back towards their original position). This corresponds for example to the case of “fanning” in the figure. Conversely, in unstable conditions, fluctuations of the transporting air appear, first in large fluctuations (looping in the figure), then with more and more turbulence leading to a large dispersion and cone (not shown in the above figure).

In a first estimation, it can be expected that *σ*_y_ and *σ*_z_ are proportional to *x* which means that the polluting plume is conical. This is due to a dimensional analysis since *x* seems to be the only fundamental length in the problem. Also, the wind induces turbulence and, in a second approach, the relation between the dispersions and *x* is no longer linear. The following curves (Turner, 1994) show this dependency (figure F-S5-3), but are focused over a large *x* (from 100 m to 20 km) since the paper was written for the description of industrial pollution. However, they are based on outdoor experiments conducted for a distance comprised between 50 and 800 meters ((Barad, 1958; Gifford, 1961; Turner, 1994) and references therein).

**Figure F-S5-2:**
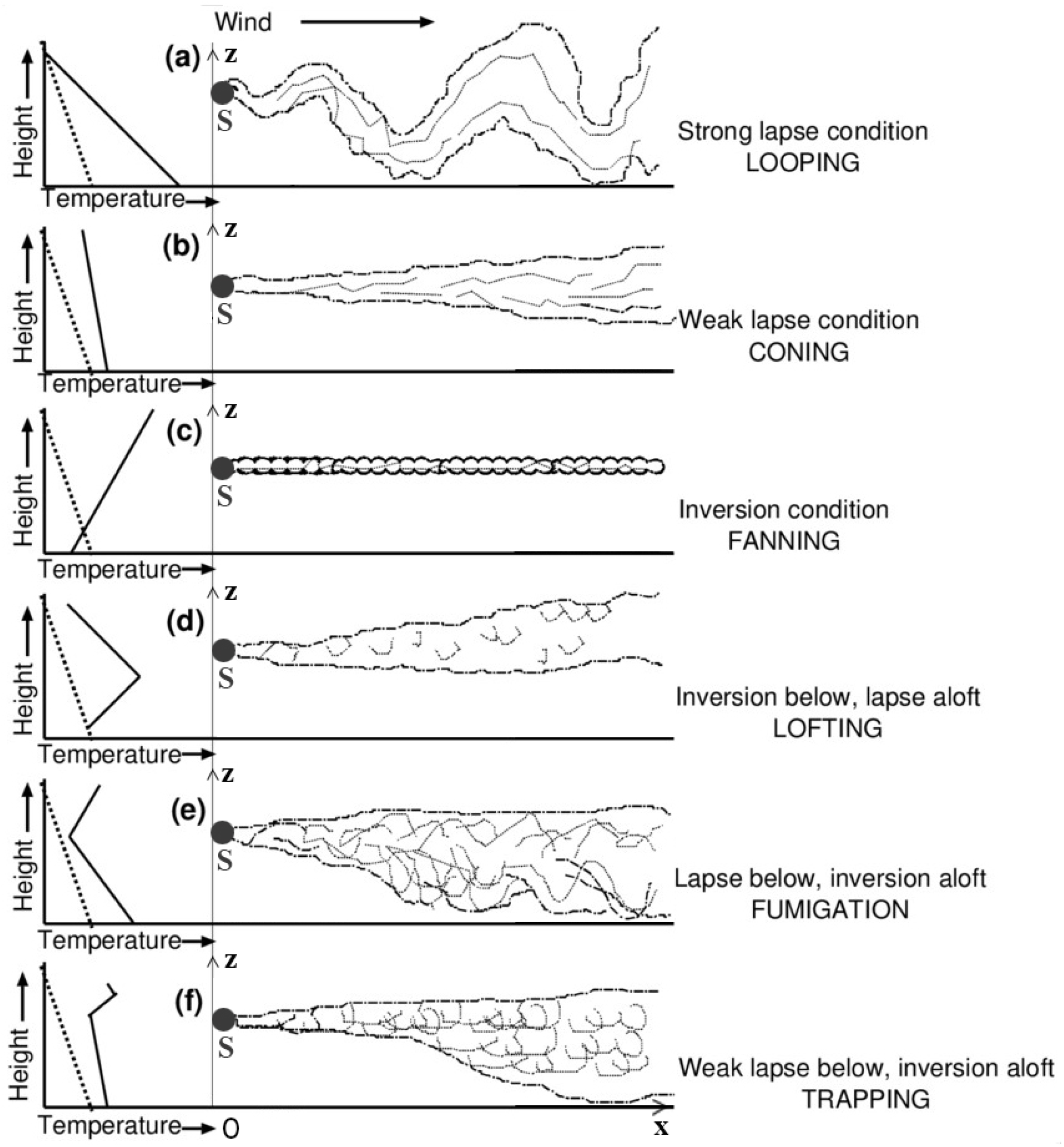
different kind of plumes following atmospheric conditions, especially lapse rate. The dotted line for the temperature is the dry adiabatic lapse rate, letters in minuscule do not refer to the stability although (a) is equivalent to A very unstable conditions. (b) refers to moderately stable or unstable conditions, cases from (c) to (d) refer to inversion, either from the ground (radiation inversion) or aloft.

**Figure F-S5-3:**
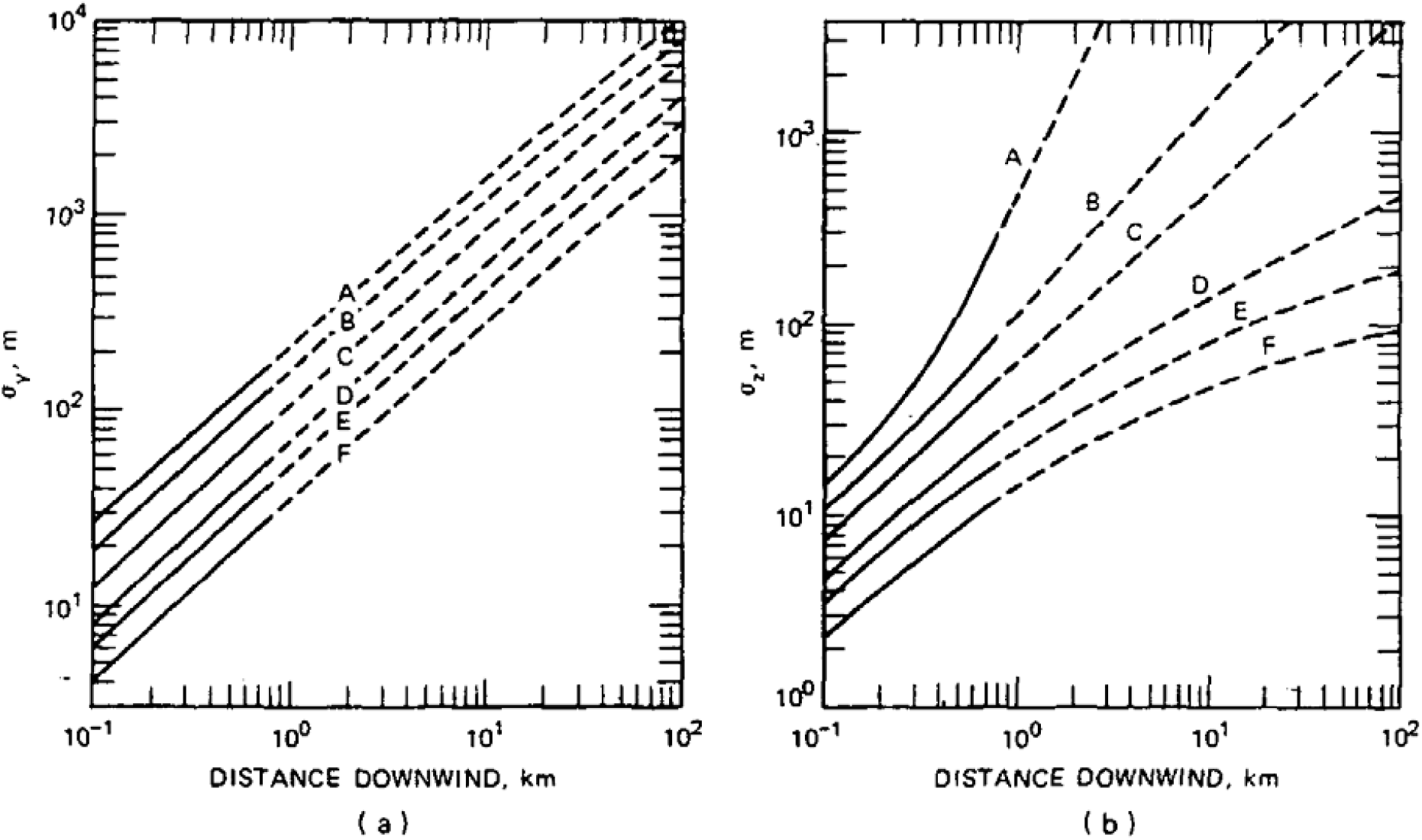
horizontal and vertical dispersion length for Gaussian plumes (Turner, 1994).

In any case, at short distances, it appears that the experimental measurements reveal a quasi-linear dependence. For small *x* (20 m to 1 km), *σ*_*y*_ and *σ*_*z*_ can be approximated as linear in *x* and the extrapolation of the above curves yields the following table:

**Table T5-2:** linear variation of dispersion lengths with distance downwind.

|  | A | B | C | D | E | F |
| --- | --- | --- | --- | --- | --- | --- |
| $\sigma_y/x$ | 0.276 | 0.199 | 0.136 | 0.085 | 0.065 | 0.042 |
| $\sigma_z/x$ | 0.2 | 0.097 | 0.074 | 0.051 | 0.034 | 0.023 |

Note that *σ*_*y*_, essentially due to the fluctuations of the wind direction, is greater than *σ*_*z*_ but as stated above we are mainly interested in *σ*_*d*_. Taking a value of the height of dispersion *H* equal to *σ*_*d*_ clearly minimizes its real value and therefore, doing so leads to an over-estimation of the outdoor risk which goes clearly in the direction of the main conclusions of the main paper.

However, figure F-S-3 given above and widely used in the literature does not cover all the situations that a plume can encounter. They are adapted to the “coning” case but not for vertical dispersion in special cases referenced as “fanning”, “fumigation” or “trapping” which can occur in cases of extremely stable inversion. Lateral dispersion may always be described by a Gaussian but in these cases there is hardly a vertical dispersion above the height of the inversion, if any, and its prediction is exceedingly difficult. In these cases, we will assume following Turner (Turner, 1994) that dispersion does not occur above this height. We choose a constant value for *H* equal to *z*_*e*_, the mean height of an emitter, which again clearly maximizes the evaluated risk outdoor.

#### Buoyancy effects

An infector expels polluted air at a temperature near 32 °C while the external temperature may be lower, say 15 °C. The puff (cough or sneeze) or the plume (speaking, breathing, or singing) are at the very beginning turbulent, emitted in a cone with an apex angle about 24°. Part of the droplets are ejected out of the plume by falling or turbulences. The smallest few ones, too small for falling on the ground, are carried away by the wind in a secondary horizontal plume; the largest droplets which have not enough time to evaporate, deposit on the ground. The remaining plume is raised by buoyancy, but this is less and less efficient because it cools down due to external air mixing. Figure displays this behavior.

**Figure F-S5-4:**
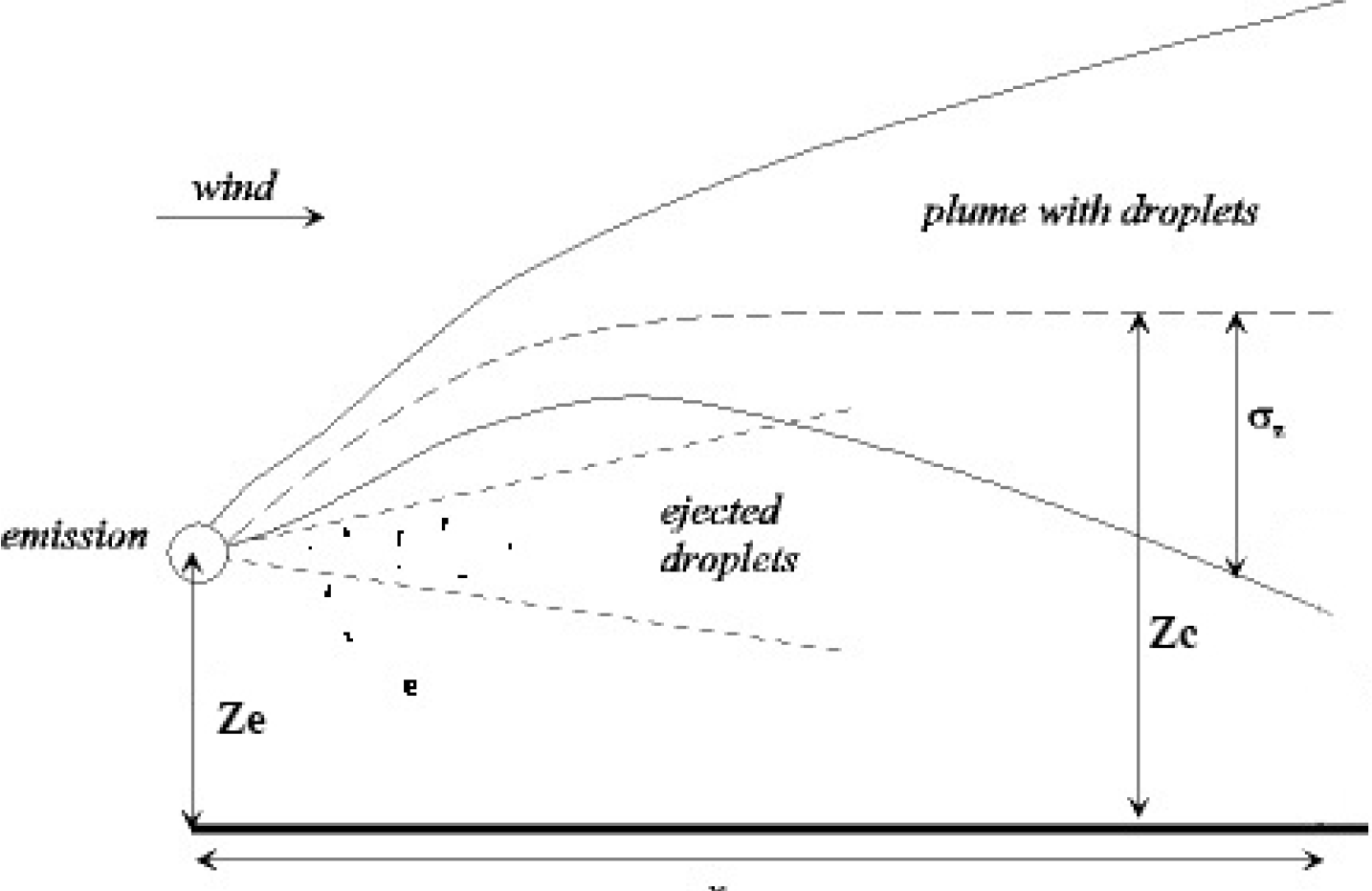
overview of a plume with buoyancy.

In a stable atmosphere the vertical dispersion remains small and outdoor the plume will eventually rise sufficiently high to pass over an exposed person. This is not the case for the secondary plume of small, ejected particles. Indoor buoyancy can bring the aerosol particles to the inlet of recirculated air.

Note therefore that the phenomena of buoyancy, which has not been taken into account in the main paper, can increase the probability of being infected indoor while most often reducing it outdoor.

